# Molecular and clinical characterization of a founder mutation causing G6PC3 deficiency

**DOI:** 10.1101/2024.05.13.24307299

**Authors:** Xin Zhen, Michael J Betti, Meltem Ece Kars, Andrew Patterson, Edgar Alejandro Medina-Torres, Selma Cecilia Scheffler Mendoza, Diana Andrea Herrera Sánchez, Gabriela Lopez-Herrera, Yevgeniya Svyryd, Osvaldo M. Mutchinick, Eric Gamazon, Jeffrey C Rathmell, Yuval Itan, Janet Markle, Patricia O’Farrill Romanillos, Saul Oswaldo Lugo-Reyes, Ruben Martinez-Barricarte

**Author notes:** Equal contribution (Saul Oswaldo Lugo-Reyes, Ruben Martinez-Barricarte). Correspondence to: Saul Oswaldo Lugo-Reyes and Ruben Martinez-Barricarte.

## Abstract

**Background:** G6PC3 deficiency is a rare genetic disorder that causes syndromic congenital neutropenia. It is driven by the intracellular accumulation of a metabolite named 1,5-anhydroglucitol-6-phosphate (1,5-AG6P) that inhibits glycolysis. Patients display heterogeneous extra-hematological manifestations, contributing to delayed diagnosis.

**Objective:** The *G6PC3* c.210delC variant has been identified in patients of Mexican origin. We set out to study the origin and functional consequence of this mutation. Furthermore, we sought to characterize the clinical phenotypes caused by it.

**Methods:** Using whole-genome sequencing data, we conducted haplotype analysis to estimate the age of this allele and traced its ancestral origin. We examined how this mutation affected G6PC3 protein expression and performed extracellular flux assays on patient-derived cells to characterize how this mutation impacts glycolysis. Finally, we compared the clinical presentations of patients with the c.210delC mutation relative to other G6PC3 deficient patients published to date.

**Results:** Based on the length of haplotypes shared amongst ten carriers of the *G6PC3* c.210delC mutation, we estimated that this variant originated in a common ancestor of indigenous American origin. The mutation causes a frameshift that introduces a premature stop codon, leading to a complete loss of G6PC3 protein expression. When treated with 1,5-anhydroglucitol (1,5-AG), the precursor to 1,5-AG6P, patient-derived cells exhibited markedly reduced engagement of glycolysis. Clinically, c.210delC carriers display all the clinical features of syndromic severe congenital neutropenia type 4 observed in prior reports of G6PC3 deficiency.

**Conclusion:** The *G6PC3* c.210delC is a loss-of-function mutation that arose from a founder effect in the indigenous Mexican population. These findings may facilitate the diagnosis of additional patients in this geographical area. Moreover, the *in vitro* 1,5-AG-dependent functional assay used in our study could be employed to assess the pathogenicity of additional *G6PC3* variants.

## INTRODUCTION

Human glucose-6-phosphatase catalytic subunit 3 (G6PC3) deficiency is an autosomal recessive disorder first identified by Boztug *et al.* in 2009 as a cause of syndromic severe congenital neutropenia (SCN)^1^. In addition to accelerated apoptosis, neutrophils from patients show impaired chemotaxis and bactericidal activity^2,3^. More than 110 patients have been described in the literature, and the clinical spectrum of G6PC3 deficiency continues to expand. Aside from neutropenia, the most frequently observed features include cardiac defects, urogenital malformations, prominent superficial veins, and endocrine abnormalities^4–6^. However, around 20% of reported cases of G6PC3 deficiency manifest as a non-syndromic form of SCN^4,7,8^. Moreover, instances of cyclic neutropenia have been observed sporadically in patients^9,10^. The phenotypic heterogeneity, coupled with the rarity of this disorder, contributes to delays in diagnosis and treatment.

In practice, it is often necessary to rule out several potential diagnoses before suspecting a genetic defect in patients with G6PC3 deficiency, and the process of identifying disease-causing variants through genetic testing afterward is intricate^11^. As most publicly available genetic databases were built using samples from individuals of European descent, alleles specific to other populations may not be well represented in these datasets^12,13^. Thus, variants observed in patients from underrepresented ancestries are more prone to being classified as variants of uncertain significance (VUS), adding further complexity in achieving a prompt diagnosis^14^. Furthermore, the absence of an effective functional test to measure the deleteriousness of novel *G6PC3* variants leads to additional challenges in establishing disease causality. Here, we describe and functionally characterize a founder mutation in *G6PC3* observed in patients from Mexico and of indigenous American ancestry.

## RESULTS

### The *G6PC3* c.210delC variant found in Mexico originated from a founder effect

Among all the mutations causing G6PC3 deficiency, a few are only found in specific ethnic groups, implying founder effects^6^. For instance, the c.210delC variant has been reported in 13 G6PC3 deficient patients, who are either homozygous or compound heterozygous for this variant ^4,15–17^. Among them, 12 patients are of Mexican descent, while another patient is included from the North American Severe Chronic Neutropenia International Registry^15^. These observations led us to interrogate whether a founder effect causes the recurrence of this mutation or if it is a mutational hotspot. To study this, we recruited four patients from central Mexico who are homozygous for this variant and were born to unrelated, non-consanguineous parents (Fig.1A-B). We collected genomic DNA from these four patients plus six of their heterozygous healthy parents. We performed whole-genome sequencing (WGS) followed by haplotype analysis on the region of chromosome 17 surrounding *G6PC3*. Since samples were unavailable from 2 of the patients’ fathers, these two carrier haplotypes were inferred computationally from the mother and patient haplotypes. Our analysis revealed a shared haplotype segment surrounding the mutation site among all carriers (Fig.1C). The length of this shared haplotype suggests that the mutation originated 26 generations ago (95% confidence interval, 2.3-50.5) in a common ancestor. This analysis indicates that the *G6PC3* c.210delC single-nucleotide deletion is a founder mutation in Mexico.

**Figure 1:**
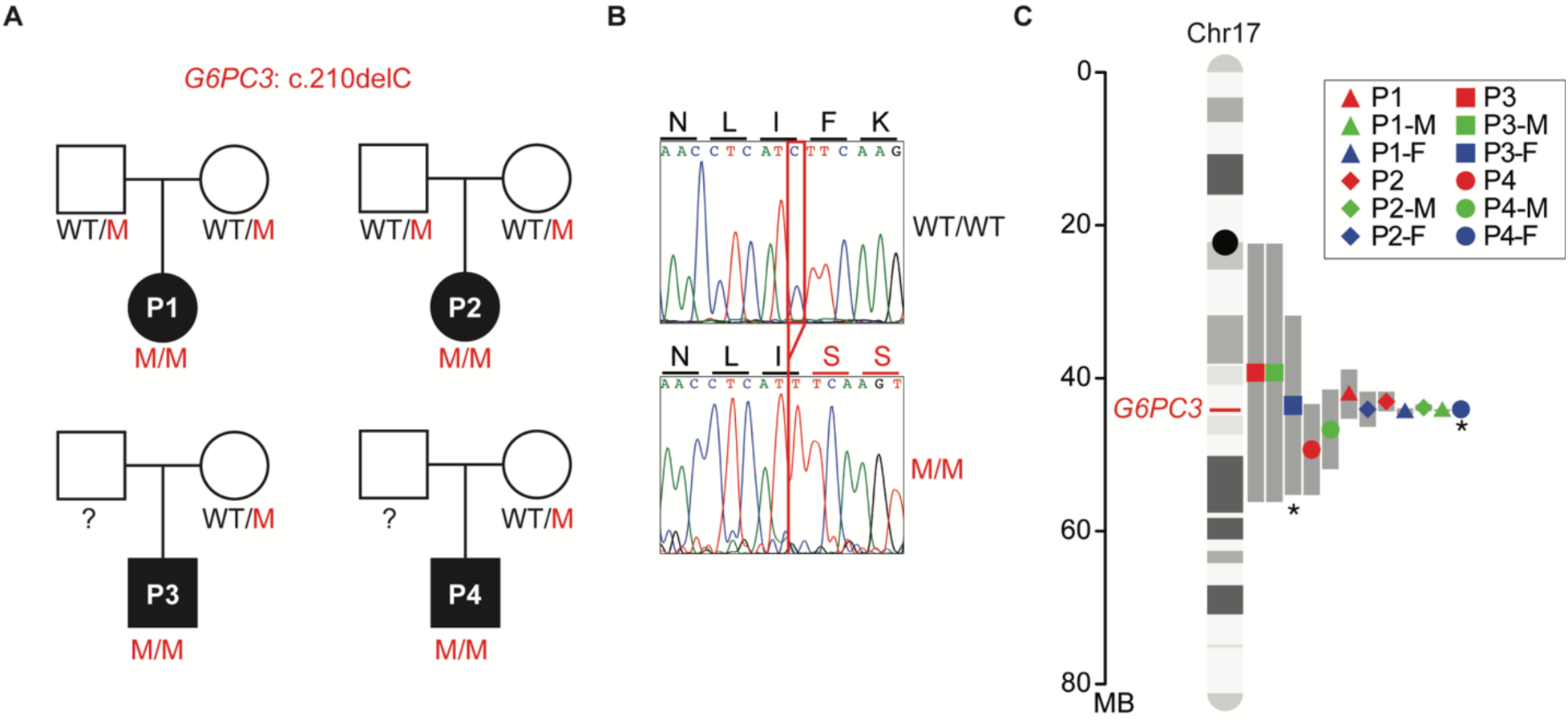
A common ancestor in patients with the *G6PC3* c.210delC mutation. **A)** Familial segregation of the mutation in four unrelated, non-consanguineous families. WT: wild type; M: mutant. **B)** Sanger sequencing results of healthy control and patient 1 (P1) in the region spanning the *G6PC3* c.210delC mutation. Amino acid changes resulting from the mutation are annotated above the graphs. **C)** Age estimation for the *G6PC3* c.210delC variant, based on the lengths of shared ancestral haplotype blocks upstream and downstream of the G6PC3 stop-gain allele (marked in red on chr17). The computationally inferred haplotypes from Father 3 (P3-F) and Father 4 (P4-F) are denoted with an asterisk.

### The *G6PC3* c.210delC mutation is of Indigenous American origin

Based on the assumption that each generation interval is between 20 and 25 years, the estimated age of the *G6PC3* c.210delC variant is 520-650 years. This allele age raised the question of whether the shared haplotype originated from native American ancestry or was introduced to Mexico by Europeans. To investigate this, we conducted principal component analysis (PCA) with whole genome sequencing (WGS) data of 10 carriers, using reference populations from the combined 1000 Genomes Project (1KGP) and Human Genome Diversity Project (HGDP) dataset. Ancestry inference analysis on the whole genome and chromosome 17 showed that the carriers of this mutation cluster closely with the indigenous American populations from HGDP (Fig.2A-B). Moreover, we used 2,343 deeply sequenced reference samples of individuals of European, African, and Ad Mixed American Ancestry from the 1KGP+HGDP dataset to perform local ancestry estimation across chr17. Our analysis indicates that the chromosomal region containing the *G6PC3* c.210delC mutation (chr17: 44,071,175) is of American origin (Fig.2C). Overall, our analysis shows that the mutation c.210delC originated in the indigenous Mexican population.

**Figure 2:**
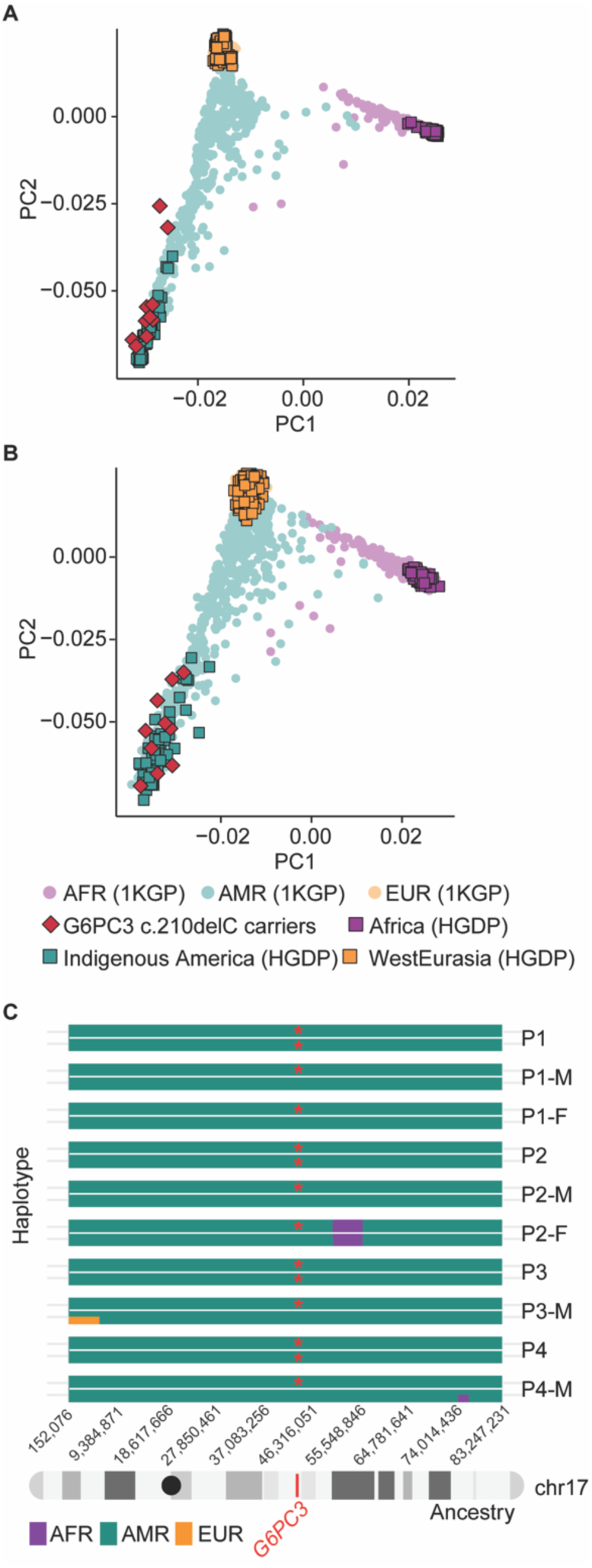
The *G6PC3* c.210delC mutation is estimated to be of indigenous American ancestry. Principal component analysis (PCA) of ancestry on **A)** the whole genome for c.210delC mutation carriers, using reference genomes from the combined 1000 Genomes Project (1kGP) and Human Genome Diversity Project (HGDP) dataset and **B)** for chromosome 17. **C)** Local ancestry estimates across carriers of the c.210delC variant. The mutation locus (chr17:44,071,175) is marked in red on chromosome 17. Mutated alleles are labeled with red asterisks. Abbreviations: AFR: African ancestry; AMR: admixed American ancestry; EUR: European ancestry.

### The *G6PC3* c.210delC variant results in reduced G6PC3 mRNA and complete loss of protein expression

*G6PC3* encodes a protein with nine transmembrane domains that localizes to the endoplasmic reticulum^18,19^. The c.210delC variant observed in our patients is a single-nucleotide deletion in exon 1 of *G6PC3* (Fig.3A). It is predicted to cause a shift in the reading frame after amino acid 70 (in the second transmembrane domain of the protein), thus introducing a premature stop codon 46 amino acids after the mutation site (p.F71Sfs*46) (Fig.3B). To examine the impact of this variant at mRNA and protein levels, we first transfected human embryonic kidney 293T (HEK293T) cells with plasmids containing C- or N-terminally Histidine-tagged versions of wildtype (WT) and p.F71Sfs*46 G6PC3. RT-qPCR results suggest that the mutation induces a moderate reduction in *G6PC3* mRNA expression (Fig.3C). Western blotting of whole cell lysate with an anti-His-tag antibody yielded a 10kDa band in the cells transfected with N-terminal tagged p.F71Sfs*46 G6PC3, aligning with the anticipated molecular weight of a truncated mutant protein (Fig.3D). This predicted mutant protein would not retain enzymatic activity as it lacks the active site of G6PC3, which is composed of amino acids R79, H114, and H167^19^ (Fig. 3B). By analyzing the C-terminal His-tagged mutant G6PC3, we showed that there is no translation re-initiation after the premature termination codon (Fig.3D). To gain additional insights in a more physiologically relevant context, we utilized Epstein-Barr Virus immortalized B (EBV-B) cells derived from two patients and healthy controls. We observed markedly reduced levels of *G6PC3* mRNA, potentially due to nonsense-mediated mRNA decay (Fig.3E). Furthermore, using membrane protein fractions extracted from EBV-B cells for western blotting and using a polyclonal antibody with epitope spanning the N-terminal region of human G6PC3, we showed that neither the WT nor the mutant size protein was expressed in cells from the patients (Fig.3F). Altogether, our data demonstrate that the *G6PC3* c.210delC (p.F71Sfs*46) variant disrupts mRNA and protein levels, causing a complete loss of expression.

**Figure 3:**
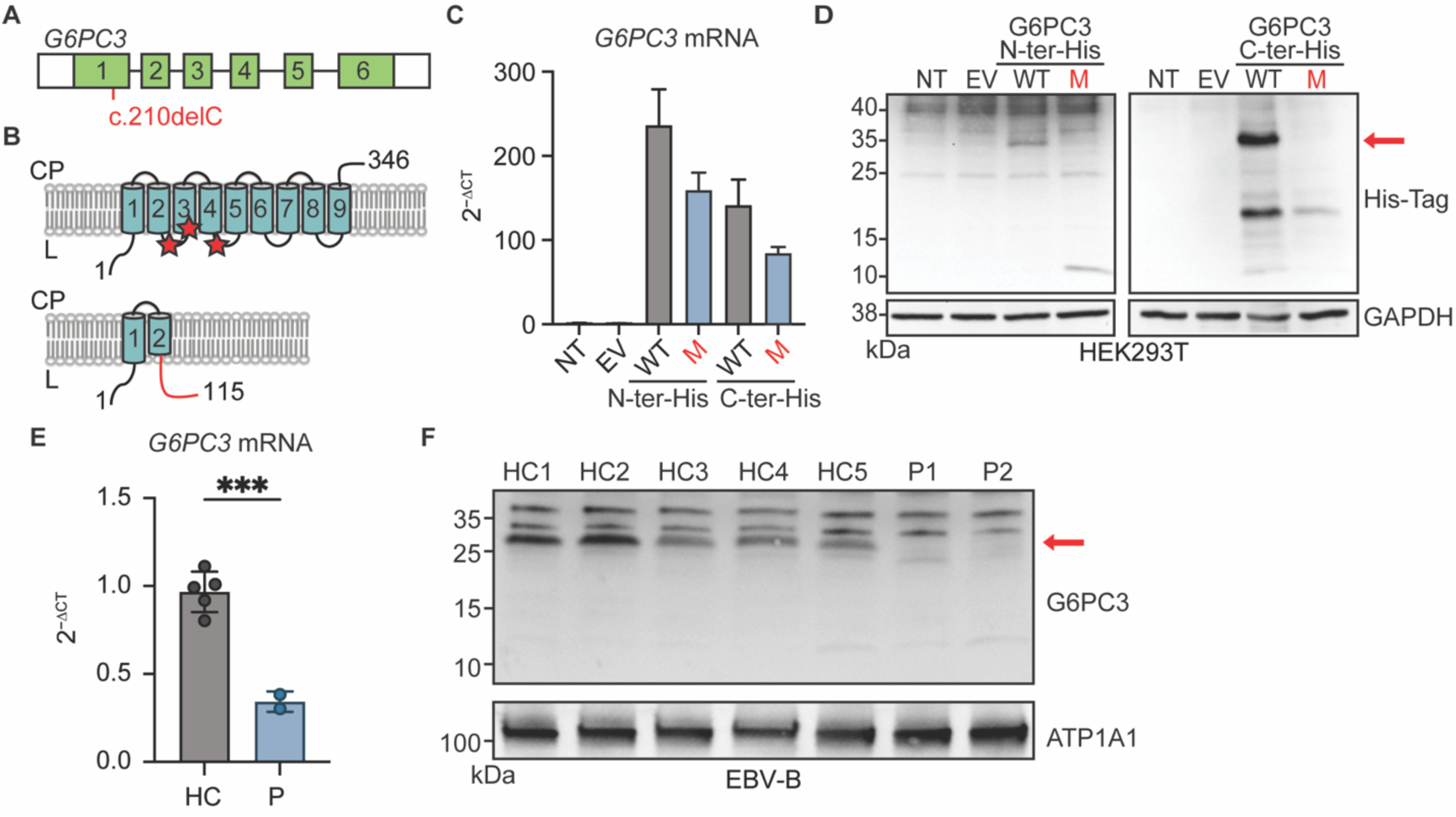
The *G6PC3* c.210delC mutation leads to a complete loss of protein expression. **A)** Cartoon of the *G6PC3* gene in which each exon is represented with a box. The mutation is indicated in red. **B)** Upper panel: Schematic representation of the G6PC3 protein structure with nine transmembrane domains in the endoplasmic reticulum. Red stars indicate the residues that compose the active site. Lower panel: the predicted consequence of the c.210delC (p.F71Sfs*46) mutation. The out-of-frame sequence resulting from the premature stop codon is indicated in red. CP: cytoplasm; L: lumen. **C)** *G6PC3* mRNA expression by RT-qPCR results of HEK293T cells either non-transfected (NT), transfected with the empty vector (EV), or plasmids encoding wildtype (WT) or F71Sfs*46 (M) G6PC3 with a 6xHis tag inserted at either the N- or C-terminal. Data are displayed as 2^−ΔCt^ after normalization to *GUS* control gene expression. n=3. **D)** Western blot of transfected HEK293T cell lysates using an anti-His Tag antibody. GAPDH was used as a loading control. **E)** RT-qPCR analysis for *G6PC3* mRNA expression in EBV-B cells from 2 patients and 5 healthy controls. Data are presented as mean ± SD. Statistical analysis was performed using Student’s t-test. ***p≤ 0.001. **F)** G6PC3 protein expression by western blot in membrane protein fraction of EBV-B cell lysates. ATP1A1 was used as a membrane protein loading control.

### Cells from patients with the *G6PC3* c.210delC variant show abolished G6PC3 enzymatic function

G6PC3 is a metabolite-repair enzyme involved in the hydrolysis of 1,5-anhydroglucitol-6-phosphate (1,5-AG6P), which is the phosphorylated form of a food-derived polyol named 1,5-anhydroglucitol (1,5-AG)^20^. When a functional G6PC3 is absent, 1,5-AG6P can accumulate to a concentration that inhibits hexokinase activity^20^. Accumulation of 1,5-AG6P thereby impairs glycolysis since hexokinase mediates the rate-limiting first step of the glycolytic pathway^21^. This mechanism leads to neutropenia since neutrophils rely heavily on glycolysis to fulfill their energetic needs^22^. To directly assess the metabolic consequence of the absence of G6PC3 caused by the c.210delC variant, we used patient-derived EBV-B cells to determine if they have defective hexokinase activity in mediating glycolysis. EBV-B cells express all four isoforms of hexokinase (I, II, III, and ADP-glucokinase) identified in mammalian cells, thus permitting us to test the functional impact of the G6PC3 deficiency (Fig. S1). EBV-B cells from patients and healthy controls were treated with either 1,5-AG or glycolytic inhibitor 2-Deoxy-D-Glucose (2-DG) for five days. Then, we quantified the glycolytic activities of these cells through measurements of extracellular acidification rate (ECAR). Patient EBV-B cells exhibited similar levels of glycolytic activity to cells derived from healthy controls, whether untreated or treated with 2-DG. However, patient cells showed significantly impaired glycolysis rate and glycolytic capacity following the 1,5-AG treatment, while healthy control cells remained unaffected (Fig. 4A-C). These results illustrate the metabolic disturbance in cells from patients with the *G6PC3* c.210delC variant due to the absence of G6PC3, establishing ECAR measurements in EBV-B cells as a means to examine the functional consequences of *G6PC3* mutations.

**Figure 4:**
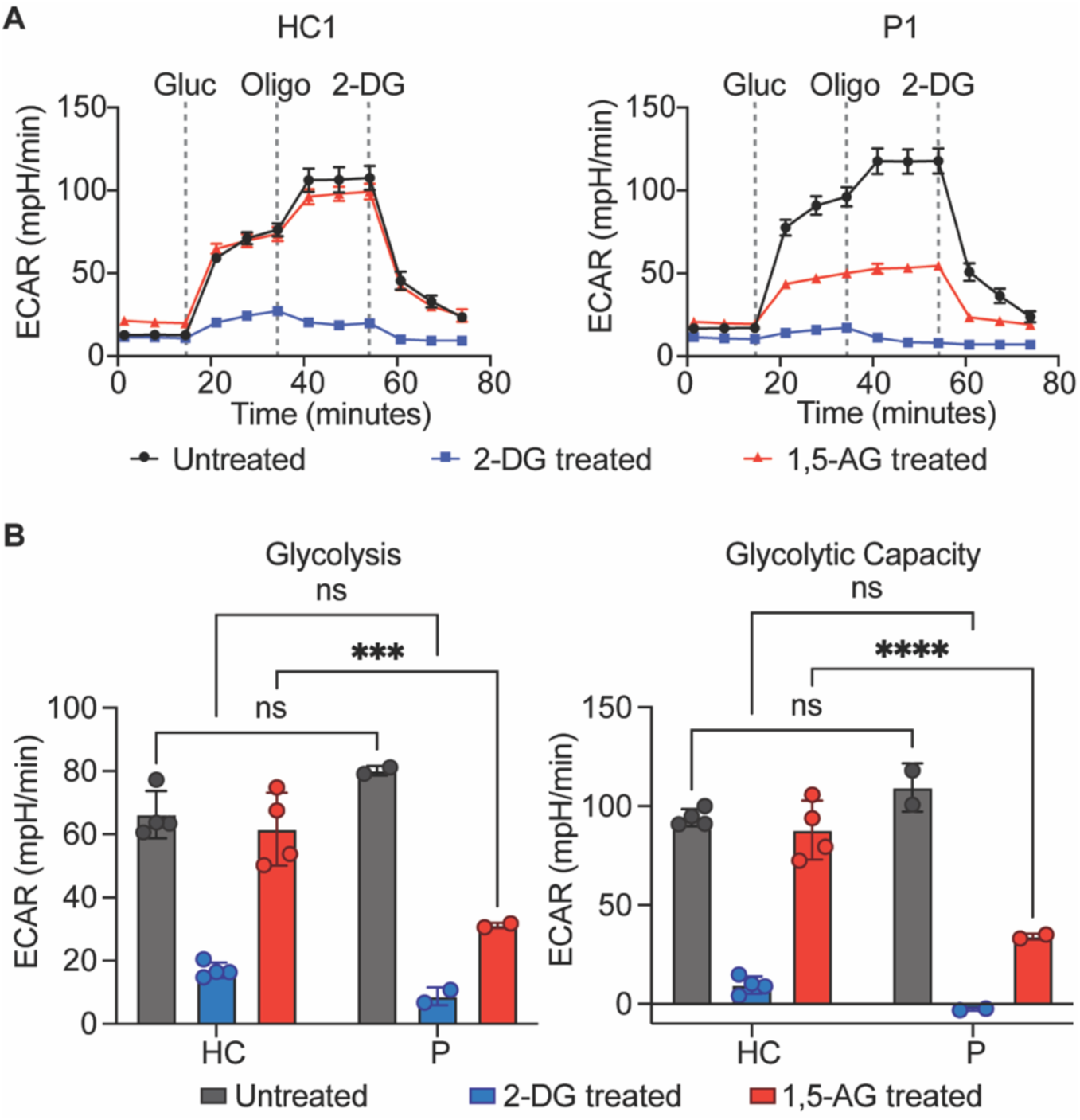
Impaired glycolysis in cells from G6PC3 deficient patients. **A)** Measurement of extracellular acidification rate (ECAR) in response to glucose (gluc), ATP synthase inhibitor oligomycin (oligo), and glycolytic inhibitor 2-deoxy-glucose (2-DG) in EBV-B cells from patients and healthy controls pretreated with 2-DG or 1,5-AG. Representative time courses of a patient (P1) and healthy control (HC1) are shown. **B)** Quantification of glycolysis rate and glycolytic capacity in EBV-B cells from four healthy controls and two patients. Data are presented as mean ± SD and represent three independent experiments. Statistical analysis was performed using two-way ANOVA with Šidák correction. ***p≤ 0.001, ****p≤ 0.0001.

### Patients with the *G6PC3* c.210delC variant show a clinical profile similar to other G6PC3-deficient patients

As all patients with the G6PC3 c.210delC variant are from the same geographical area, we evaluated whether they exhibit any characteristic in their clinical presentation that may differentiate them from the rest of the reported G6PC3 deficient patients denoting some environmental aspects of the disease. To this end, we collected the clinical information from all published cases of G6PC3 deficiency to compare the frequency of appearance of nine prominent clinical features between patients with and without the *G6PC3* c.210delC mutation (n=14, some published, some unpublished) (Fig. 5). None of these patients showed isolated neutropenia but present with features of syndromic severe congenital neutropenia including extra-hematological abnormalities. Except for hepatosplenomegaly, all other features of G6PC3 deficiency have been observed in these patients. We also noticed that patients with the c.210delC variant display a higher occurrence of thrombocytopenia, endocrine abnormalities, and hearing loss. Although this may imply a specific characteristic of this group of patients, it might also reflect some variability in standard clinical testing. Overall, our analysis shows that patients who are carriers of the *G6PC3* c.210delC mutation display all main clinical characteristics described in G6PC3 deficiency, indicating that the mutation may be the main driver of their disease.

**Figure 5:**
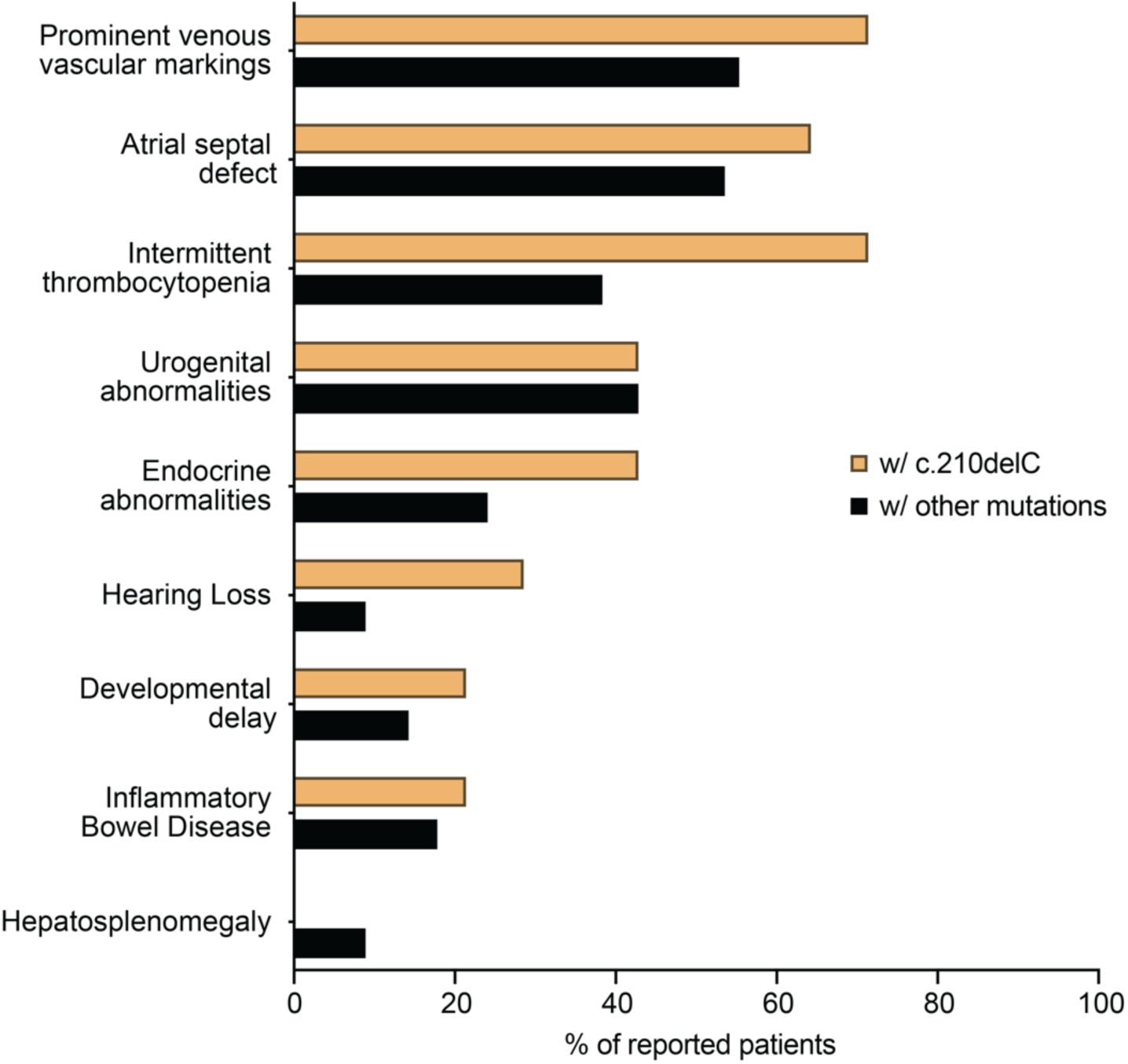
Frequencies of the most prominent clinical features observed in G6PC3 deficient patients with the c.210delC mutation (n=14) and those with other mutations (n=112).

## DISCUSSION

G6PC3 deficiency is a rare genetic disorder with a broad phenotypic spectrum, posing difficulties for timely diagnosis. The differential diagnosis process can be particularly complicated for patients with non-syndromic neutropenia or less frequently observed clinical features^23–25^. Mutations observed in G6PC3 deficient patients spread across all six exons of the gene^6^. Interestingly, the prevalence of several G6PC3 alleles varies significantly amongst different ethnic groups. These include the p.Phe44Ser mutation of Pakistani origin and the p.W73X mutation in patients from the Dominican Republic^26,27^. Here, through haplotype analysis and ancestry inference, we demonstrated that the *G6PC3* c.210delC variant was recurrently observed in Mexico due to a founder effect, and it is of native American origin. We also reviewed the signs and symptoms in patients with this variant, concluding that they closely resemble those observed in other reported patients. These findings may facilitate targeted testing of patients from this region with unexplained congenital neutropenia.

The mechanisms responsible for the phenotypic variability in G6CP3 deficiency remain elusive thus far^6,8,28^. It has been hypothesized that these variations might be correlated with the residual G6PC3 enzymatic activity resulting from some mutations^29^. To assess the functional impact of mutations, previous studies measured the capacity of mutant G6PC3 in mediating hydrolysis of glucose-6-phosphate (G6P) using microsomes isolated from transfected yeast or COS-1 cells^1,29^. However, it has been recently established that the primary physiological role of G6PC3 is not to dephosphorylate G6P into glucose and phosphate. Instead, the molecular mechanism underlying neutrophil dysfunction observed in G6PC3 deficient patients is associated with the accumulation of 1,5-AG6P^20,30^. This highlights the need for a 1,5-AG-dependent functional test to assess the pathogenicity of G6PC3 variants. Using Seahorse extracellular flux assays, we have shown that EBV-B cells from our patients fail to eliminate 1,5-AG6P, leading to defective glycolytic activity, indicating that the *G6PC3* c.210delC variant disrupts the metabolite repair activity of G6PC3. One limitation is that we utilize immortalized cells rather than primary cells from patients to illustrate this effect. Nevertheless, this *in vitro* assay could still be employed in future studies to determine the impact of *G6PC3* mutations in additional patients, which would aid in establishing potential genotype-phenotype correlations for this disease, as well as predicting the pathogenicity of variants of uncertain significance (VUS).

In conclusion, this study identified the *G6PC3* c.210delC allele as a founder mutation that abolishes protein expression and function. These findings may help expedite the diagnosis of G6PC3 deficiency, especially in the Mexican population. As previous reports suggest that G6PC3 deficiency can lead to death from severe infections when neutropenia is left untreated, prompt diagnosis and provision of treatments are critical^31,32^. Importantly, empagliflozin, an SGLT2 inhibitor frequently used to treat type 2 diabetes, has successfully resolved neutrophil defects in patients with G6PC3 deficiency by lowering the 1,5-AG blood concentrations^30,33,34^. Along with the use of this highly effective, safe, and easy-to-take oral alternative to granulocyte-colony-stimulating factor (G-CSF) injections, early disease diagnosis may improve outcomes of G6PC3 deficient patients^35,36^.

## Data Availability

All data produced in the present work are contained in the manuscript

## ACKNOWLEDGMENT

We would like to thank the patients and their families for supporting the study. RMB was funded in part by the National Institute of Allergy and Infectious Diseases (R21AI171466 and R01AI168210) and the National Cancer Institute (R01CA269217).

## MATERIALS AND METHODS

### Estimating *G6PC3* allele age

Blood-derived DNA collected from 10 carriers of the *G6PC3* c.210delC allele (two trios and two mother-child pairs) were sequenced using Illumina short-read sequencing. The affected chromosome (chr17) was sequenced at an average read depth of 10.92X across all ten individuals. Raw reads were aligned to human reference genome build GRCh38/hg38 using Burrows-Wheeler Aligner ^37^ (0.7.17). PCR duplicates were subsequently identified in the aligned reads, and base quality scores were recalibrated following the GATK Best Practices Workflow^38^. Pre-called GVCFs from these six individuals underwent subsequent joint variant calling using the HaplotypeCaller^39^ function from GATK (4.2.0.0). Finally, variant quality scores were recalibrated for both SNPs and indels.

The jointly called VCF was pruned using VCFtools^40^ (0.1.15) and VcfFilter from the BIOPET suite^41^ (0.2). After removing variants with any missingness across the six individuals, those with a minor allele count of less than 3, a quality score of less than 30, or a minimum read depth of less than 20 were also removed.

Using this pruned variant set, the affected chromosome (chr17) was phased using SHAPEIT4^42^ (4.2.0), with 3,202 1000 Genomes samples sequenced at 30X^43^ serving as the reference panel. The phased VCF was converted to hap/sample format using bcftools^44^ (1.12). Using R (4.0.3), the hap file was reformatted as a 13-column text file containing the chr21 coordinate and phased variant calls for each of the 12 phased chromosomes (2 per individual) at that site. Phased variant calls could then be manually spot-checked for concordance with the aligned reads using the IGV genome browser^45^ (2.9.4). As genetic data from the fathers were unavailable in two of the four pedigrees, these carrier haplotypes were inferred from the mother and proband haplotypes using a custom Python script. The lengths in centimorgans (cM) of the chr17 chromosomal arms upstream and downstream of the mutation site were calculated, and these respective lengths were used as inputs for the Mutation_age_estimation.R script developed by Gandolfo et al. ^46^. A confidence coefficient of 0.95 without chance sharing correction was used, assuming a correlated genealogy.

### Ancestry inference using PCA

To determine the genetic ancestries of the *G6PC3* c.210delC carriers, we combined the variant calls from 10 samples with those from 2,343 reference samples with African, American, and European ancestries from the harmonized HGDP + 1KGP dataset^47^. Variant filtering was performed according to Hardy-Weinberg equilibrium (*P* < 1×10^−6^) and missingness using PLINK v.1.9^48^. Autosomal variants with a minor allele frequency >1% were retained and pruned for linkage disequilibrium (*r*^2^ = 0.2). PCA was conducted using 157,601 variants with smartpca implemented in EIGENSOFT version 8.0.0^49^. Eigenvectors were calculated using 2,343 reference samples, and the 10 *G6PC3* c.210delC carriers were projected onto these eigenvectors. A second PCA was performed using 8,032 variants located on chr17.

### Estimating local ancestry

Using the phase_common_static function from SHAPEIT (v5.5.1)^50^, variant calls from the ten sequenced carriers of the *G6PC3* c.210delC allele were phased together with 2,343 deeply sequenced reference samples (788 of European ancestry, 1,003 of African origin, and 552 of American ancestry) from the gnomAD v3.1.2 HGDP + 1KG callset^51^ (https://gnomad.broadinstitute.org/downloads#v3-hgdp-1kg). After phasing, RFMix (v2.03-r0)^52^ was used to estimate local ancestry across chr17 for the ten carriers of the *G6PC3* c.210delC allele.

### Establishment of Epstein-Barr Virus (EBV)-Immortalized B Cell Lines

For generation of EBV-B cell lines derived from patients and healthy control individuals, purified B cells were immortalized with EBV as previously reported^53^.

### Cell culture

EBV-B cell lines were cultured in RPMI 1640 Medium without glucose (Gibco, Carlsbad, CA), supplemented with 5mM glucose (Gibco) and 10% fetal bovine serum (FBS) (Corning, Corning, NY). HEK293T cells (ATCC; CRL-3216) were cultured in DMEM medium (Sigma-Aldrich, St. Louis, MO) supplemented with 10% FBS (Corning).

### Plasmid cloning and site-directed mutagenesis

RNA was extracted from healthy control EBV-B cells using the RNeasy Plus Mini Kit (Qiagen, Venlo, The Netherlands). RNA was reverse transcribed using the Verso cDNA Synthesis Kit (ThermoFisher Scientific, Waltham, MA). Primer sequences used to generate the full-length cDNA of *G6PC3* with a His-tag right next to the start or stop codon of cDNA that encodes the G6PC3 protein were as follows: N-ter His-tag Forward: 5’-ATG CAT CAC CAT CAC CAT CAC ATG GAG TCC ACG CTG G-3’; N-ter His-tag Reverse: 5’-TCA GGA AGA GTG GAT GGG-3’; C-ter His-tag Forward: 5’-ATG GAG TCC ACG CTG G-3’; C-ter His-tag Reverse: 5’-TCA GTG ATG GTG ATG GTG ATG GGA AGA GTG GAT GGG C-3’. The products were TA-cloned into a pcDNA3.1 plasmid vector using the pcDNA 3.1/V5-His TOPO TA Expression Kit (ThermoFisher Scientific) according to the manufacturer’s protocol. Plasmid DNAs were purified from bacterial clones with a Miniprep kit (ThermoFisher Scientific). Then, constructs carrying the 210delC mutant allele were generated by site-directed mutagenesis with primers: Forward: 5’-CTC AAC CTC ATT TCA AGT GGT T-3’; Reverse: 5’-AAC CAC TTG AAA TGA GGT TGA G-3’. In brief, after the PCR reaction using PfuUltra II Fusion High-fidelity DNA Polymerase (Agilent, Santa Clara, CA), 1uL DpnI (NEB, Ipswich, MA) was added followed with incubation at 37C for 3 hours. Mutagenesis was validated by Sanger sequencing.

### HEK293T cell transfection

In 6-well plates, HEK293T cells were transfected in 2 mL media with 2.5ug of pcDNA3.1 empty vector, N or C-terminal His-tagged WT *G6PC3*, or N- or C-terminal His-tagged 210delC mutant constructs using Lipofectamine 3000 Transfection Reagent (ThermoFisher Scientific) according to the manufacturer’s protocol for 48 hours.

### *G6PC3* mRNA detection by RT-qPCR

Total RNA was extracted from transfected HEK293T cells and EBV-B cells using the RNeasy Kit (Qiagen, Venlo, The Netherlands). The RT-qPCR was performed using the Luna Universal One-Step RT-qPCR Kit (NEB) on the CFX96 RT-qPCR detection system (Bio-Rad, Hercules, CA). The following primers were used to amplify the cDNA of *G6PC3*: Forward: 5’-TCA AGT GGT TTC TTT TTG GAG-’3; Reverse: 5’-ATC ATG CAG TGT CCA GAA G-’3. The *G6PC3* mRNA expression level in each sample was normalized to the expression level of the *GUS* gene transcript.

### G6PC3 detection by western blotting analysis

Total cell lysates were prepared from transfected HEK293T cells with RIPA lysis buffer (150 mM NaCl, 50 mM TRIS-HCl pH 8.0, 1 mM EDTA, 0.5% sodium deoxycholate, 1% NP-40, 0.1% SDS), supplemented with protease inhibitor cocktail (ThermoFisher Scientific). Protein concentrations were quantified with DC Protein Assay (Bio-Rad). Lysates mixed with Laemmli loading buffer were incubated at 100°C for 5 minutes and subjected to electrophoresis in a 10% protein gel.

The membrane protein fraction of EBV-B cells was obtained with the Mem-PER Plus Membrane Protein Extraction Kit (ThermoFisher Scientific). The membrane protein extracts were mixed with loading buffer, incubated at 37°C for 30 minutes, and subjected to electrophoresis in a precast 4%-20% gradient gel (Bio-Rad).

The following antibodies were used for Western blotting: HRP-conjugated Anti-His tag (Biolegend, 652503, 1:5000), Anti-G6PC3 (Invitrogen, PA5-109749, 1:1000), Anti-ATP1A1 (Invitrogen, MA5-32184, 1:1000), HRP-conjugated Anti-GAPDH (Proteintech, HRP-60004, 1:1000), Anti-HK1 (Cell signaling, 2804, 1:1000), Anti-HK2 (Proteintech, 22029-1-AP, 1:3000), Anti-HK3 (Invitrogen, PA5-29304, 1:1000), and Anti-ADPGK (Proteintech, 15639-1-AP, 1:1000). Unconjugated antibodies were detected by incubation with HRP Goat Anti-mouse IgG secondary antibody (Sigma-Aldrich, AP127P, 1:5000) or HRP Goat Anti-Rabbit IgG secondary antibody (Sigma-Aldrich, AP156P, 1:5000).

### Extracellular flux assay

EBV-B cells were pretreated with 2mM of 2-DG (Cayman Chemical, Ann Arbor, MI) or 1,5-AG (Cayman Chemical) for five days prior to the glycolysis stress assay. Cells were counted and resuspended in Agilent Seahorse XF RPMI 1640 medium supplemented with 2mM glutamine, with 2mM of 2-DG or 1,5-AG added to corresponding conditioned cells. 150,000 live cells/well were plated. ECAR was measured on a Seahorse XFe 96 bioanalyzer using the glycolysis stress test with sequential injections of 100mM glucose, 15µM oligomycin, and 500mM 2-DG. Glycolysis rate was quantified as the change in ECAR before and after addition of glucose. Glycolytic capacity was calculated as the difference between maximal ECAR reached following the oligomycin injection and the non-glycolytic acidification prior to addition of glucose.

### Ethics statement

This study was conducted in accordance with the Helsinki Declaration, with written informed consent obtained from the patients and their families. Approval for this study was obtained from the Vanderbilt University Medical Center Institutional Review Board (IRB), Nashville, USA.

## Supplementary material

**Supplementary Figure 1:**
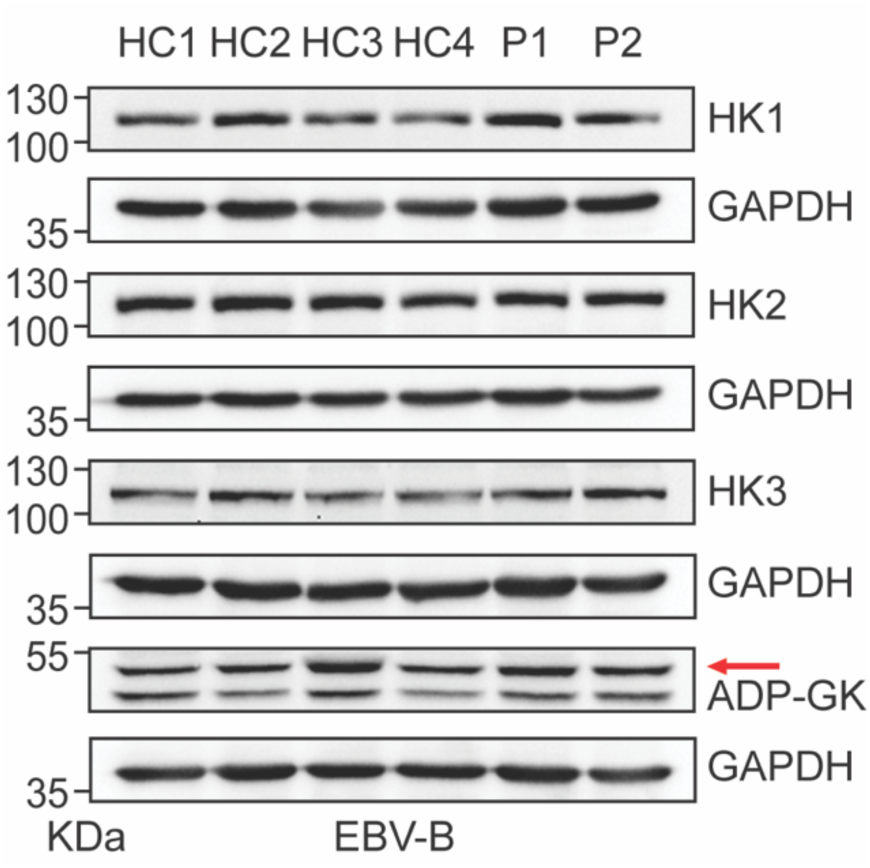
Hexokinase expression in EBV-B cells. Western blot expression of the four isoforms of hexokinase in EBV-B cells. GAPDH was used as the loading control.

## Notes

### Competing Interest Statement

The authors have declared no competing interest.

### Author Declarations

Institutional Review Board (IRB) of Vanderbilt University Medical Center gave ethical approval for this work

## REFERENCE

1. Boztug, K. et al. A Syndrome with Congenital Neutropenia and Mutations in G6PC3. New England Journal of Medicine 360, 32–43 (2009).

2. McKinney, C. et al. Metabolic abnormalities in G6PC3-deficient human neutrophils result in severe functional defects. Blood Adv 4, 5888 (2020).

3. Dai, R. et al. Altered Functions of Neutrophils in Two Chinese Patients With Severe Congenital Neutropenia Type 4 Caused by G6PC3 Mutations. Front Immunol 12, (2021).

4. Velez-Tirado, N. et al. Severe congenital neutropenia due to G6PC3 deficiency: Case series of five patients and literature review. Scand J Immunol 95, (2022).

5. Maroufi, S. F. et al. Novel G6PC3 Mutations in Patients with Congenital Neutropenia: Case Reports and Review of the Literature. Endocr Metab Immune Disord Drug Targets 21, 1660–1668 (2021).

6. Banka, S. & Newman, W. G. A clinical and molecular review of ubiquitous glucose-6-phosphatase deficiency caused by G6PC3 mutations. Orphanet J Rare Dis 8, (2013).

7. Khera, S., Pramanik, S. K. & Patnaik, S. K. A Novel Mutation in G6PC3 Gene Associated Non-syndromic Severe Congenital Neutropenia. Indian Pediatr 57, 574–575 (2020).

8. Banka, S., Wynn, R., Byers, H., Arkwright, P. D. & Newman, W. G. G6PC3 mutations cause non-syndromic severe congenital neutropenia. Mol Genet Metab 108, 138–141 (2013).

9. Bruni, C. M., de la Rua, W., Sadre, S. Y., Nestor, J. M. & Ahmed, R. A Novel Pathogenic Variant of G6PC3 Gene Presenting As Cyclic Neutropenia in a Pediatric Patient. Blood 138, 4191 (2021).

10. Alangari, A. A., Alsultan, A., Osman, M. E., Anazi, S. & Alkuraya, F. S. A novel homozygous mutation in G6PC3 presenting as cyclic neutropenia and severe congenital neutropenia in the same family. J Clin Immunol 33, 1403–1406 (2013).

11. Kiykim, A. et al. G6PC3 deficiency: Primary immune deficiency beyond just neutropenia. J Pediatr Hematol Oncol 37, 616–622 (2015).

12. Gudmundsson, S. et al. Variant interpretation using population databases: Lessons from gnomAD. Hum Mutat 43, 1012 (2022).

13. Sirugo, G., Williams, S. M. & Tishkoff, S. A. The Missing Diversity in Human Genetic Studies. Cell 177, 26 (2019).

14. Chen, E. et al. Rates and Classification of Variants of Uncertain Significance in Hereditary Disease Genetic Testing. JAMA Netw Open 6, E2339571 (2023).

15. Xia, J. et al. Prevalence of mutations in ELANE, GFI1, HAX1, SBDS, WAS and G6PC3 in patients with severe congenital neutropenia. Br J Haematol 147, 535–542 (2009).

16. Boztug, K. et al. Extended spectrum of human glucose-6-phosphatase catalytic subunit 3 deficiency: novel genotypes and phenotypic variability in severe congenital neutropenia. J Pediatr 160, (2012).

17. López-Rodríguez, L. et al. Severe Congenital Neutropenia Type 4: A Rare Disease Harboring a G6pc3 Gene Pathogenic Variant Particular to the Mexican Population. Rev Invest Clin 74, 328–339 (2022).

18. Guionie, O., Clottes, E., Stafford, K. & Burchell, A. Identification and characterisation of a new human glucose-6-phosphatase isoform. FEBS Lett 551, 159–164 (2003).

19. Shieh, J. J., Pan, C. J., Mansfield, B. C. & Chou, J. Y. A Glucose-6-phosphate Hydrolase, Widely Expressed Outside the Liver, Can Explain Age-dependent Resolution of Hypoglycemia in Glycogen Storage Disease Type Ia. Journal of Biological Chemistry 278, 47098–47103 (2003).

20. Veiga-da-Cunha, M. et al. Failure to eliminate a phosphorylated glucose analog leads to neutropenia in patients with G6PT and G6PC3 deficiency. Proc Natl Acad Sci U S A 116, 1241–1250 (2019).

21. Roberts, D. J. & Miyamoto, S. Hexokinase II integrates energy metabolism and cellular protection: Akting on mitochondria and TORCing to autophagy. Cell Death & Differentiation 2015 22:2 22, 248–257 (2014).

22. Jeon, J. H., Hong, C. W., Kim, E. Y. & Lee, J. M. Current Understanding on the Metabolism of Neutrophils. Immune Netw 20, 1–13 (2020).

23. Moradian, N. et al. Severe congenital neutropenia due to G6PC3 deficiency: early and delayed phenotype of a patient. *Allergy*, Asthma and Clinical Immunology 19, 1–8 (2023).

24. Notarangelo, L. D. et al. Severe congenital neutropenia due to G6PC3 deficiency: Early and delayed phenotype in two patients with two novel mutations. Ital J Pediatr 40, 1–6 (2014).

25. Yildirmak, Z. Y., Ozcelik, G., Ozagari, A. A., Genc, D. B. & Onay, H. Amyloidosis in a Patient with Congenital Neutropenia because of G6PC3 Deficiency. J Pediatr Hematol Oncol 44, E431–E433 (2022).

26. Glasser, C. L. et al. Phenotypic Heterogeneity of Neutropenia and Gastrointestinal Illness Associated with G6PC3 Founder Mutation. J Pediatr Hematol Oncol 38, e243–e247 (2016).

27. Smith, B. N. et al. Phenotypic heterogeneity and evidence of a founder effect associated with G6PC3 mutations in patients with severe congenital neutropenia. Br J Haematol 158, 146–149 (2012).

28. Banka, S., Wynn, R. & Newman, W. G. Variability of bone marrow morphology in G6PC3 mutations: Is there a genotype–phenotype correlation or age-dependent relationship? Am J Hematol 86, 235–237 (2011).

29. Lin, S. R., Pan, C. J., Mansfield, B. C. & Chou, J. Y. Functional analysis of mutations in a severe congenital neutropenia syndrome caused by glucose-6-phosphatase-β deficiency. Mol Genet Metab 114, 41–45 (2015).

30. Boulanger, C. et al. Successful use of empagliflozin to treat neutropenia in two G6PC3-deficient children: Impact of a mutation in SGLT5. J Inherit Metab Dis 45, 759–768 (2022).

31. Alizadeh, Z. et al. Two Cases of Syndromic Neutropenia with a Report of Novel Mutation in G6PC3. Iran J Allergy Asthma Immunol 10, 227–230 (2011).

32. Fernandez, B. A. et al. Adult siblings with homozygous G6PC3 mutations expand our understanding of the severe congenital neutropenia type 4 (SCN4) phenotype. BMC Med Genet 13, 111 (2012).

33. Lédeczi, Z., Pittner, R., Kriván, G., Kardon, T. & Legeza, B. Empagliflozin restores neutropenia and neutrophil dysfunction in a young patient with severe congenital neutropenia type 4. J Allergy Clin Immunol Pract 11, 344–346.e1 (2023).

34. Hiwarkar, P. et al. SLGT2 Inhibitor Rescues Myelopoiesis in G6PC3 Deficiency. J Clin Immunol 42, 1653–1659 (2022).

35. Veiga-da-Cunha, M., Wortmann, S. B., Grünert, S. C. & Van Schaftingen, E. Treatment of the Neutropenia Associated with GSD1b and G6PC3 Deficiency with SGLT2 Inhibitors. Diagnostics 13, (2023).

36. Dale, D. C., Bolyard, A. A. & Makaryan, V. The promise of novel treatments for severe chronic neutropenia. Expert Rev Hematol 16, 1025–1033 (2023).

37. Li, H. & Durbin, R. Fast and accurate short read alignment with Burrows-Wheeler transform. Bioinformatics 25, 1754–1760 (2009).

38. McKenna, A. et al. The Genome Analysis Toolkit: A MapReduce framework for analyzing next-generation DNA sequencing data. Genome Res 20, 1297–1303 (2010).

39. Poplin, R., et al. Scaling accurate genetic variant discovery to tens of thousands of samples. bioRxiv 201178 (2018) doi:10.1101/201178.

40. Danecek, P. et al. The variant call format and VCFtools. Bioinformatics 27, 2156 (2011).

41. Hof, P. V. T. et al. BIOPET: Towards scalable, maintainable, user-friendly, robust and flexible NGS data analysis pipelines. Proceedings - 2017 17th IEEE/ACM International Symposium on Cluster, Cloud and Grid Computing, CCGRID 2017 823–829 (2017) doi:10.1109/CCGRID.2017.59.

42. Delaneau, O., Zagury, J. F., Robinson, M. R., Marchini, J. L. & Dermitzakis, E. T. Accurate, scalable and integrative haplotype estimation. Nature Communications 2019 10:1 10, 1–10 (2019).

43. Byrska-Bishop, M. et al. High-coverage whole-genome sequencing of the expanded 1000 Genomes Project cohort including 602 trios. Cell 185, 3426–3440.e19 (2022).

44. Danecek, P. et al. Twelve years of SAMtools and BCFtools. Gigascience 10, 1–4 (2021).

45. Thorvaldsdóttir, H., Robinson, J. T. & Mesirov, J. P. Integrative Genomics Viewer (IGV): high-performance genomics data visualization and exploration. Brief Bioinform 14, 178– 192 (2013).

46. Gandolfo, L. C., Bahlo, M. & Speed, T. P. Dating rare mutations from small samples with dense marker data. Genetics 197, 1315–1327 (2014).

47. Koenig, Z., et al. A harmonized public resource of deeply sequenced diverse human genomes. bioRxiv 2023.01.23.525248 (2024) doi:10.1101/2023.01.23.525248.

48. Purcell, S. et al. PLINK: a tool set for whole-genome association and population-based linkage analyses. Am J Hum Genet 81, 559–575 (2007).

49. Price, A. L. et al. Principal components analysis corrects for stratification in genome-wide association studies. Nature Genetics 2006 38:8 38, 904–909 (2006).

50. Hofmeister, R. J., Ribeiro, D. M., Rubinacci, S. & Delaneau, O. Accurate rare variant phasing of whole-genome and whole-exome sequencing data in the UK Biobank. Nature Genetics 2023 55:7 55, 1243–1249 (2023).

51. Koenig, Z. et al. A harmonized public resource of deeply sequenced diverse human genomes. bioRxiv 2023.01.23.525248 (2023) doi:10.1101/2023.01.23.525248.

52. Maples, B. K., Gravel, S., Kenny, E. E. & Bustamante, C. D. RFMix: A Discriminative Modeling Approach for Rapid and Robust Local-Ancestry Inference. Am J Hum Genet 93, 278 (2013).

53. Tosato, G. & Cohen, J. I. Generation of Epstein-Barr Virus (EBV)–Immortalized B Cell Lines. Curr Protoc Immunol 76, 7.22.1–7.22.4 (2007).

